# Alternative Approaches for Monitoring and Evaluation of Lymphatic Filariasis Following Mass Drug Treatment with Ivermectin, Diethylcarbamazine and Albendazole in East New Britain Province, Papua New Guinea

**DOI:** 10.1101/2024.04.03.24305242

**Authors:** Krufinta Bun, Benedict Mode, Melinda Susapu, Catherine Bjerum, Michael Payne, Daniel Tisch, Makoto Sekihara, Emanuele Giorgi, Gary J. Weil, Peter U Fischer, Leanne Robinson, Moses Laman, Christopher L. King

## Abstract

**Background:** WHO recommends two annual rounds of mass drug administration (MDA) with ivermectin, diethylcarbamazine, and albendazole (IDA) for lymphatic filariasis (LF) elimination in treatment naïve areas that are not co-endemic for onchocerciasis such as Papua New Guinea (PNG). Whether two rounds of MDA are necessary or sufficient and the optimal sampling strategies and endpoints for stopping MDA remain undefined.

**Methods and Findings:** Two cross-sectional studies were performed, one at baseline in 2019 before MDA-IDA, and 12 months post-MDA-IDA. Pre-MDA, we selected 49 sentinel villages for LF in East New Britain Province (ENBP, PNG) and randomly sampled ∼100 individuals/village of approximately equal number of children 6-9 years (N=1,906), and those ≥10 years (N=2,346) using population proportionate and purposeful sampling. LF infection was assessed by tests for circulating filarial antigenemia (CFA) and microfilariae (Mf). Children ages 6-9, 1.9% (37/1,906, range 0-21.6%) were CFA positive at baseline, and 0.3% (5/1,906; range 0-7.8%) were Mf positive. Individuals ≥10 years, 7.5% (176/2,346, range 0-52%) were CFA positive, and 2.0% (47/2,346, range 0-36%) were Mf positive. Twenty-four of 49 clusters were CFA ≥2%, and 14 had Mf prevalence ≥1%. Post-MDA (82% coverage), 47 clusters were selected based on geospatial modeling (N=4,610), of which 38 had >2% CFA compared to 24 identified at baseline. In the 24 villages evaluated pre- and post-MDA, we stratified the impact of MDA-IDA on children 6-9 and adults ≥18 years. Children had a 34% reduction in CFA prevalence and complete Mf clearance. Adults had a 39% reduction in CFA prevalence and a 96% reduction in Mf prevalence. Post-MDAx1 showed no villages that were Mf positive in two of four districts.

**Conclusions:** Geospatial modeling was more effective in sampling high-risk sites for LF than population-proportional sampling. The low LF prevalence in children and slight reduction of CFA prevalence limits its utility as a biomarker for LF elimination in children. A single round of MDA with IDA with high coverage was sufficient to reach elimination targets in villages with low baseline LF prevalence. Areas with higher baseline prevalence will require additional rounds of MDA, but this could be targeted to smaller evaluation units to reduce cost.

**Trial registration:** This study is registered at Clinicaltrials.gov under the number NCT04124250

**Author Summary:** *Why was this study done?:*

- WHO has targeted lymphatic filariasis (LF) for global elimination as a public health problem using mass drug administration (MDA) as the primary intervention strategy.
- The WHO recently modified recommendations for MDA of LF with a combination of three co-administered drugs: ivermectin, diethylcarbamazine, and albendazole. This study examined the impact of one round of MDA on LF infection parameters in Papua New Guinea that had not previously received MDA for LF and examined new methodologies for monitoring and surveillance.

*What did the researcher do and find?:*

- Before MDA, we randomly sampled sentinel clusters (villages) using population proportional sampling of equal numbers of children 6-9 years and older children and adults using well-established LF infection parameters. Post-MDA, we selected sentinel villages using a geospatial modeling design and focused on sampling adults.
- Population-proportional sampling underestimated the overall LF infection because the infection was more common in less-densely populated rural areas. Sampling children 6-9 years of age was inefficient because of very low infection rates in this age group. Geospatial modeling was more effective than population proportional sampling for selecting areas at high risk for LF. One round of MDA with high coverage was highly effective for reducing microfilaremia prevalence to very low levels in most sampled villages, but CFA prevalence decreased less dramatically.

*What do these findings mean?:*

- Geospatial modeling and sampling adults for microfilaria are preferred methods for monitoring the impact of MDA with IDA.
- Results from this study suggest that one round of high-coverage MDA may be sufficient to interrupt LF transmission in areas with low baseline prevalence. Additional rounds of MDA can then be targeted to areas with higher LF prevalence, thus reducing program costs. This strategy requires high-quality baseline surveillance to capture the focality of LF infection and high-quality MDA. This approach may be especially useful in areas like Papua New Guinea, where MDA is logistically challenging.

## Introduction

Lymphatic filariasis (LF) is a disease caused by parasitic nematodes transmitted by mosquitoes. The infection leads to a spectrum of clinical outcomes that range from asymptomatic microfilaremia (parasite larvae circulating in the blood) to hydrocele, lymphedema, and elephantiasis [1]. Lymphatic filariasis is caused by the nematodes *Wuchereria bancrofti*, *Brugia malay*i and *Brugia timori*, of which *W. bancrofti* accounts for over 90% of infection worldwide [2]. The World Health Organization (WHO) currently estimates that 883 million people in 44 countries are at risk for LF”L/o: and that 15 million are disfigured and incapacitated by the disease [3]. The WHO started the Global Program to Eliminate Lymphatic Filariasis (GPELF) in 2000 with a goal to eliminate LF by 2020 [2], which was later revised to 2030 [4]. The elimination strategy comprises community-wide mass drug administration (MDA) in endemic areas based on administrative units that often include a mix of areas with high and low endemicity. Mass drug treatment comprises annual single-dose therapy of albendazole (ALB) combined with diethylcarbamazine (DEC) in countries not co-endemic for onchocerciasis, and ALB combined with ivermectin (IVM) in onchocerciasis co-endemic countries in sub-Saharan Africa that are not co-endemic for *Loa loa* [5]. Areas in Africa that are co-endemic for *Loa loa* and have high parasite loads should receive ALB only. The LF elimination strategy also includes morbidity management of advanced clinical cases. To interrupt transmission, the current GPELF strategy recommends MDA with a minimum effective coverage of ≥65% for up to five annual rounds of MDA [6]. This strategy has been highly successful in many LF endemic countries, resulting in 740 million people no longer requiring MDA. This amounts to a 52% reduction in the at-risk population worldwide [7]. However, this approach has been less successful in other LF endemic areas because of poor uptake of drugs in people with the highest risk of infection (e.g. adult men, migrants, and pregnant women), logistical challenges for drug delivery, and limited efficacy of the two-drug MDA regimens.

In 2017, the WHO endorsed the use of a single co-administered IVM plus DEC plus ALB ( referred to as IDA) for LF elimination in many areas outside of Africa because it was more effective than DEC plus ALB [8–13]. IDA is recommended in LF endemic settings that are eligible for drug combinations that include DEC; i) where onchocerciasis is not co-endemic, ii) that have not yet started MDA, iii) that have received fewer than four effective rounds of MDA, or iv) in areas where MDA results have been suboptimal [14]. Currently, the WHO recommends two annual rounds of MDA with IDA in eligible countries with at least 65% epidemiological coverage. More data are needed on whether two rounds are necessary or sufficient to interrupt LF transmission.

One challenge associated with MDA that is especially important in areas that distribute IDA is knowing when infection prevalence has been reduced to levels that no longer can support sustained transmission so that MDA can be stopped. While IDA is very efficient for clearing microfilariae (Mf) from the blood, circulating filarial antigen (CFA) persists because IDA sterilizes but does not kill all adult worms [9]. The current MDA stopping protocol uses transmission assessment surveys (TAS) to detect CFA in children 6-7 years of age as an indicator of recent transmission [15]. However, TAS can be insensitive to ongoing LF transmission in areas where infection prevalence is much lower in children than adults. Before proceeding to TAS, programs perform pre-TAS surveys to assess whether CFA prevalence is less than 2% or Mf prevalence is <1% in one sentinel and one spot-check site within an evaluation unit. However, the focal distribution of LF represents an important limitation to this strategy; pre-TAS surveys may miss persistent infection when only some sites are sampled in large evaluation units. Another problem with pre-TAS and TAS surveys based on the detection of CFA is that the persistence of CFA after IDA may require MDA to be continued long after Mf prevalence has been reduced to levels that cannot sustain transmission.

Cognizant of these challenges, the Task Force for Global Health (with external input) developed a new monitoring and evaluation approach to evaluate the impact of MDA with IDA. We applied this approach to East New Britain Province (ENBP), which had yet to implement province-wide MDA for LF. Here, we aim to i) assess the impact of one round of IDA on LF infection parameters, ii) provide evidence to justify adopting new elimination criteria and endpoints using IDA and iii) optimize sampling strategies to capture better the focality of LF in the population and to demonstrate the elimination of LF.

## Methods

### Study Site

East New Britain Province (ENBP, population approximately 376,566) comprises the eastern half of the island of New Britain in PNG (5° 10′ 0″ S, 151° 45′ 0″ E). It is divided into four districts: Gazelle, Pomio, Kokopo, and Rabaul (**Figure 1**, **Table 1**). Most people reside in the less mountainous northeastern part of the province. Cocoa, copra, and palm oil are the major cash crops. Rabaul and Kokopo districts are more urbanized, with more modern structures (housing with window screens), easy access to paved roads, and electricity. Gazelle and Pomio are rural districts with less infrastructure.

**Figure 1.**
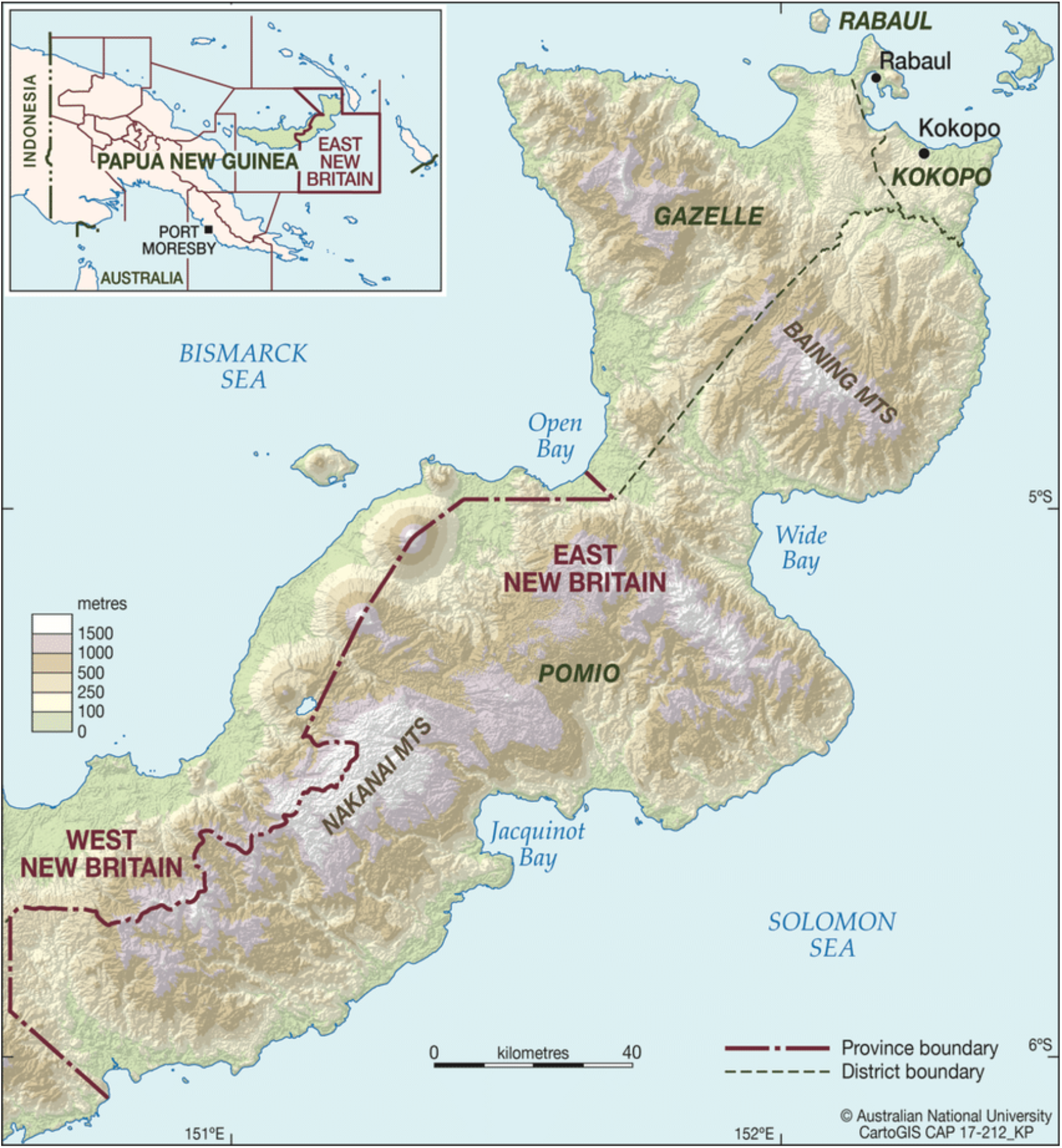
Map of East New Britain Province.

**Table 1.**
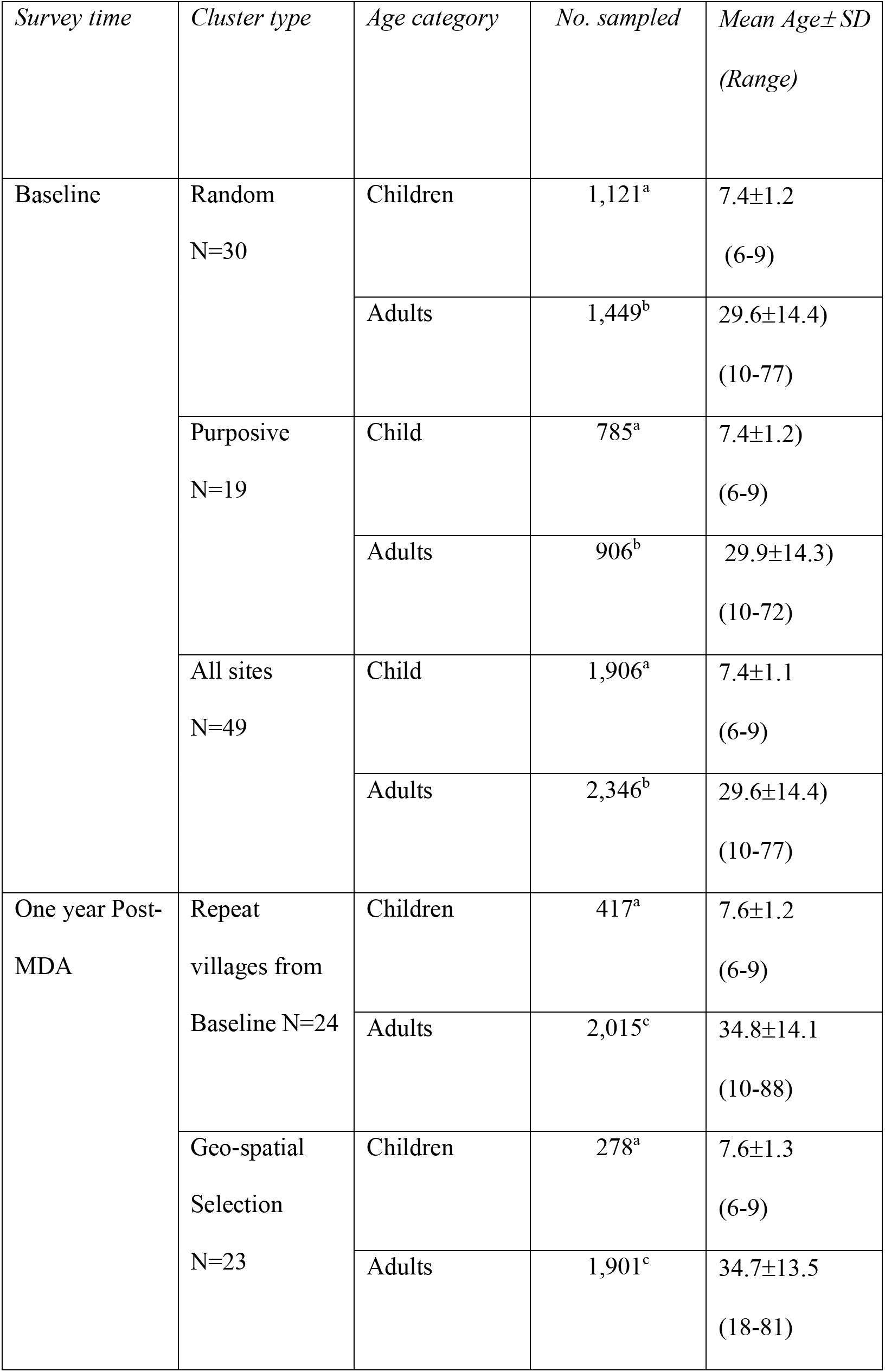
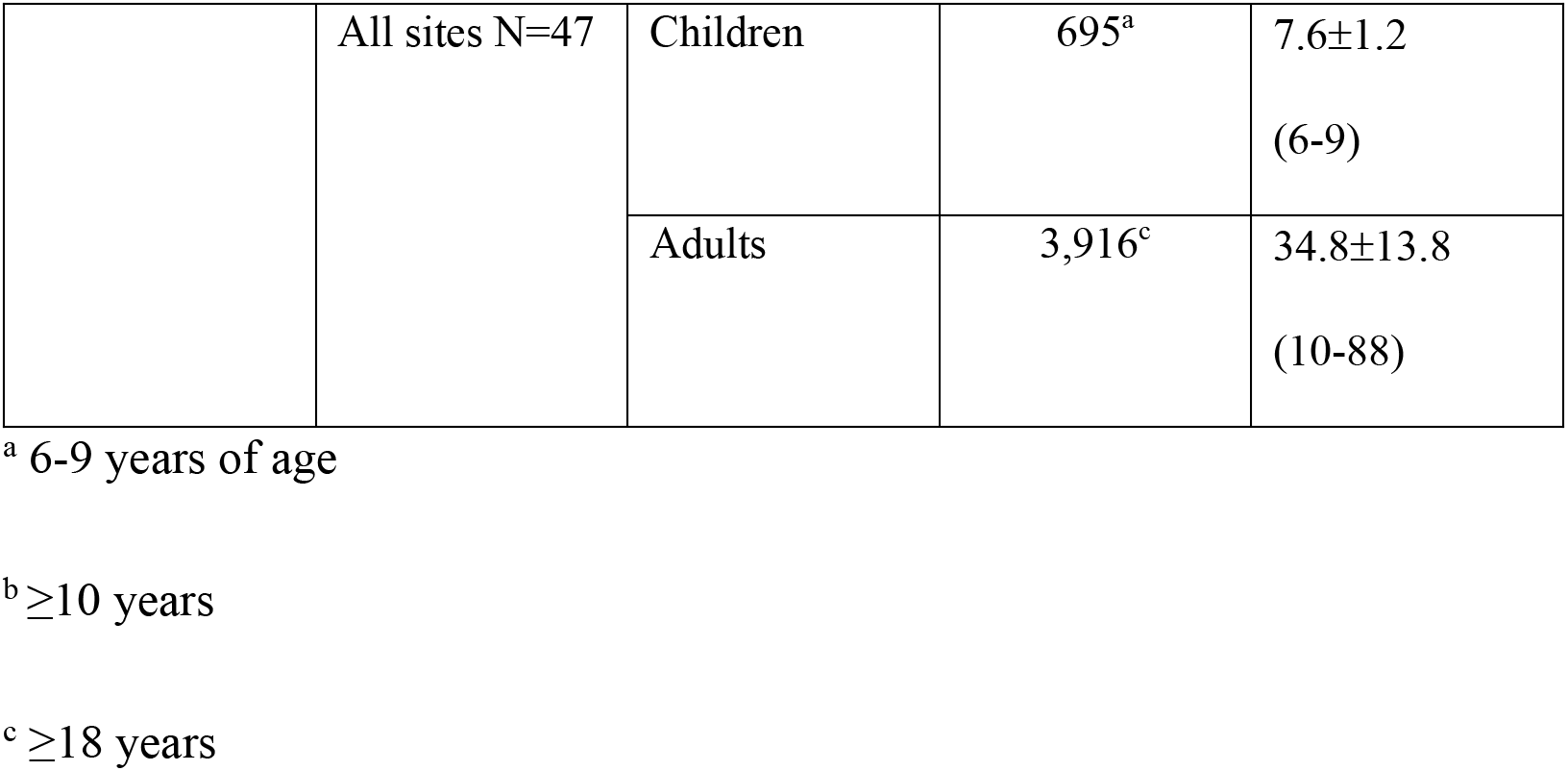
Baseline and one year post MDA-IDA impact survey characteristics.

### Ethical approval

The protocol was approved by the Medical Research Advisory Council of PNG (MRAC 19.14) and Case Western Reserve University IRB (STUDY20191141). Individual written informed consent was provided by adults aged 18 years and older. If the individuals could not read or write, a village member who could read or write served as a witness and cosigned the informed consent form. Children’s participation required the consent of their parents and documented assent for children older than 12 years of age.

### Mass Drug Administration

In November 2019 MDA with IDA (MDA-IDA) was conducted throughout the province. Before MDA, community health workers (CHW) were essential in establishing strong community engagement through meetings (known as *Toksave*), and training community distributors. MDA was distributed over 3 weeks to facilitate travel to remote areas, better capture mobile populations, and provide time for mop-up. The whole MDA process was coordinated by a highly engaged LF/NTD task force in the province that involved community leaders and other stakeholders, ensuring campaign success.

### Study design and village selection

The primary sampling unit was the village. In September and October 2019, we conducted a baseline cross-sectional survey of 30 villages in ENBP using population proportionate random sampling (PPS). Five additional villages suspected to have high LF rates were purposively selected by the ENBP provincial health LF Taskforce. Fourteen additional villages were added to improve sampling in rural areas with lower population densities and higher LF risk. The villages selected by PPS in each district (Kokopo, Rabaul, Pomio, and Gazelle) were proportionate to the population size of the districts, with Gazelle having the most selected villages and Rabaul having the least (**Table 1**).

Since the *Anopheles* mosquitos transmit LF in PNG the sample size was powered to detect a 2% CFA threshold for each of two groups; i) children aged 6-9 years old and other community members ≥10 years, and ii) during the post-MDA survey children aged 6-9 years old and other community members ages ≥18 years. The post-MDA target population and sample size were adjusted based on WHO interim guidelines of a 1% Mf threshold in adults ≥18 years with a 95% confidence interval to assess the impact of IDA (IDA Technical Report 2022). The selected households within each cluster are adapted from the WHO guidelines for evaluating coverage surveys and monitoring [16]. Fifty households were randomly selected from a starting point in the center of the community using a pen or spin bottle. Before the survey, each household within the villages was assigned a number with the help of a village recorder or leader. Household numbers were written on cut paperboard placed in a large jar, and we randomly drew numbers until 50 households were selected. If a cluster of households consisted of family groups, the cluster was considered ‘one’ household unit. All members of the 50 households were surveyed for children aged 6-9 years and individuals ≥10 years until 100 participants were reached (50 children and 50 adults). Demographic surveys recorded the number of people per household, the age and sex of individuals, the number of children ages 6-9, and the number of persons ≥10 years of age.

### Socio-economic assessment

At baseline, we compiled a socioeconomic impact (SEI) score for each village by assessing access to potable water, the use of window screens, and the level of education for each household. In calculating the SEI scores, we counted 1 point if a household had access to a water tank or pipe (0 for river/stream or public pump/well), and 1 point if window screens were used (0 for no screens). In scoring educational status, each adult (18 years or older) received a score based on their highest level of education attained; 3 points for college or vocational training, 2 points for high school graduates, 1 point for an 8th-grade education, and a score of 0 if the adult had not attended school through the 8th-grade. The composite educational score was computed by taking the average score of each adult in the household. The total SEI score for each household was obtained by adding the educational average to the water and screen scores. To get the SEI score for each village, we averaged all the households in that village. Higher SEI scores indicate higher socioeconomic status. We also computed 95% confidence intervals and ranges for village SEI scores. The SEI scores were used in the geostatistical model described below.

### Coverage survey

A cluster sampling approach was applied to determine drug coverage after one round of MDA [16]. The primary sampling unit was local level government (LLG) within each of the 4 districts, and each LLG contained wards that consisted of many villages. The Rotary Against Malaria (RAM) survey for 2016 was used as the denominator of the village population. In this survey, 8 LLGs or clusters in each of the 4 districts were selected by population proportionate sampling (PPS) which were: Gazelle Central Rural (Gazelle), Lasul Baining Rural (Gazelle), Vunadirir Toma Rural (Gazelle), Bitapaka Rural (Kokopo), Kokopo Vunamami Urban (Kokopo), East Pomio Rural (Pomio), Balanataman Rural (Rabaul) and Rabaul Urban (Rabaul). Gazelle Central Rural LLG was divided into 3 independent clusters because of the large population size relative to the rest of LLGs in ENBP. PPS then selected the villages within the LLGs for the coverage surveys. Across the LLGs, 45 villages were surveyed for drug coverage. Twenty-five of the 45 villages were in the Gazelle district, 10 in Kokopo, 5 in Pomio and 5 in Rabaul. Of the 45 villages surveyed, 15 (33.3%) were included in the baseline LF prevalence survey.

### Selection of study sites at 1-year post-MDA

The post-MDA survey used model-based geostatistics (MBG) to select 50 villages with a high probability of having a CFA prevalence >2% (Table 1). Twenty-four of the 50 selected villages had CFA >2% in the baseline survey. Approximately 100 individuals were surveyed in each village. Of the 23 newly selected villages surveyed, only adults ≥18 years were sampled because of the low infection prevalence in young children in the baseline surveys in most villages. In the 24 villages sampled pre- and post-MDA, 15 villages had CFA prevalence >2% in children 6-9 years of age at baseline. Children in these 15 villages were resampled post-MDA as follows: ∼50 children ages 6-9 and ∼50 adults ≥18 years to examine the impact of MDA on LF infection parameters in children compared to adults.

### Procedures

Fingerstick blood samples were collected during the surveys for CFA testing with Filariasis Test Strips (FTS) (Alere, Scarborough, ME, USA) according to the manufacturer’s protocol. Tests with no control line were invalid and repeated. The FTS detects a biomarker for infection with *W. bancrofti* (Wb) adult worms, and it has a high sensitivity for detecting persons with microfilaremia. In community studies in PNG, all Mf-positive individuals had positive FTS results [17]. In this study, persons with positive FTS results were tested for Mf with 60-μl thick blood smears prepared from fingerstick samples collected between 9 PM and 1 AM, as previously described [17]. Two microscopists reviewed the slides. If Mf counts were highly discordant or one reader identified Mf and the other did not, a third reader reviewed the slide.

### Data acquisition and management

The study used an electronic data capture (EDC) system using Epi Info^TM^ (Centers for Disease Control) software to capture and transfer data to a REDCap database (v11.0.3, Vanderbilt University). The data were entered by designated, trained members of the PNG research team on the day of enrollment. A parallel participant key (separate from the REDCap database and maintained at PNG Institute for Medical Research) linked study ID numbers with personal identifying information, such as name and date of birth. The participant key was not shared with Case Western Reserve University investigators or staff.

### Geospatial and Statistical analysis

The geostatistical methods used to sample the number of clusters at one-year post-MDA-IDA followed adaptive geostatistical designs [18]. The approach involved sampling or selecting locations given previous infection data. The methodological framework includes prevalence data modelled to describe the number of people infected with a particular disease or outcome, at a given location, in a specific region of interest with a vector of associated covariates. The model assumes a spatial Gaussian process to measure the outcome of interest (i.e., disease prevalence) given a set of covariates independent from each other at specific locations. The covariates in the model were environmental exposures and socio-economic impact score using the PrevMap package [19]. Records with complete spatial information correspond to 47 of the 49 clusters. The model accounts for the spatial correlation between clusters based on their proximity in space, whereby closer clusters are assumed to be more correlated than clusters further apart. The current analysis also modeled the association between locations on distinct land masses through the Euclidean distance - i.e., the ordinary “straight-line” distance. The predictions were carried out on a 1 by 1 km regular grid. Grid locations whose distance from the closest data-locations is larger than 15 km are excluded from the predictions. This is because we estimate a scale of the spatial correlation of about 15 km; hence, the data bear very little information on the prevalence at those locations. There were 748 clusters in ENBP of which we selected 49 that had the probability of exceeding 2% CFA prevalence threshold (**see Supplementary Methods**).

Descriptive statistics were calculated as frequencies and proportions for categorical variables ± 95% confidence intervals. For demographic variables, median and interquartile ranges, or mean ± SD were used. Comparisons between treatment groups for demographic and infection variables were performed using chi-square and Fisher’s exact tests (as appropriate) for categorical variables using SAS (v 9.4, SAS Institute).

## Results

The evaluation unit in PNG is the province, although the districts better capture the province’s considerable geographic and social heterogeneity. Overall, 4,252 participants were enrolled for the baseline survey and 4,611 for the one-year post-MDA-IDA survey. **Table 1** describes the population characteristics and sampling design pre- and post-MDA-IDA.

### Baseline LF infection parameters

At baseline, 4,252 participants in 49 clusters (villages) were examined for LF infection, of which 1,906 were children aged 6-9 years, and 2,346 were individuals aged ≥ 10 years (**Tables 1, 2 a and b**). Females accounted for 48% of the 6-9 years sampled and 61% of those ≥ 10 years. Among the 6-9-year-olds, 1.9 % (95% CI 1.4-2.7; 37/1,906) were CFA positive prevalence, and 0.3% (95% CI 0.09-0.61; 5/1,906) were Mf positive. Among those ≥10 years, 7.5% (95% CI 6.5-8.6; 176/2,346) were CFA positive and 2.0% (95%CI 1.5-2.7; 47/2,347) Mf positive. Circulating filarial antigen and Mf prevalence increased with age, with the highest prevalence in men between 21 and 30 (**Figure 2**). Combining all groups, twenty-four (48.9%) of the 49 villages had ≥2% CFA, and 14 met the ≥1% Mf threshold (**Supplemental Table 1A**). Based on WHO criteria, these are thresholds indicative of ongoing LF transmission. The CFA prevalence <2.0% among 6-9-year-olds would suggest that ENBP might not have ongoing LF transmission based on the WHO transmission assessment survey (TAS) of lower primary school-aged children 6-7 years of age using a 2% CFA threshold [20]. To examine this age group, we stratified the 1,906 6-9 year-old children sampled into those 6-7 years of age (N=1,075), representing 54% of children examined. Thus, there would be approximately 24,000 6-7 year of age children in ENBP (Table 1; 45,188 x 0.54 = 24,400). Using a community-based TAS algorithm of a population size of ∼20,000, the sample size would be ∼1,500 in 30 clusters, like the current baseline survey. Based on all 6-7 year-old children surveyed in 49 clusters, CFA prevalence was 2.1% (95% CI 1.69-2.40, 22/1075), and none were Mf positive. In the 30 community-based clusters selected by PPS, the CFA prevalence in 6-7-year-olds was 0.6% (95% CI 0.2-1.6%, 4/625), below the 2% CFA threshold, although this sample size is smaller than WHO recommendation, yet the upper 95% CI is <2%. Thus, using the standard TAS survey might have led to an erroneous conclusion that ENBP would not need MDA.

**Figure 2.**
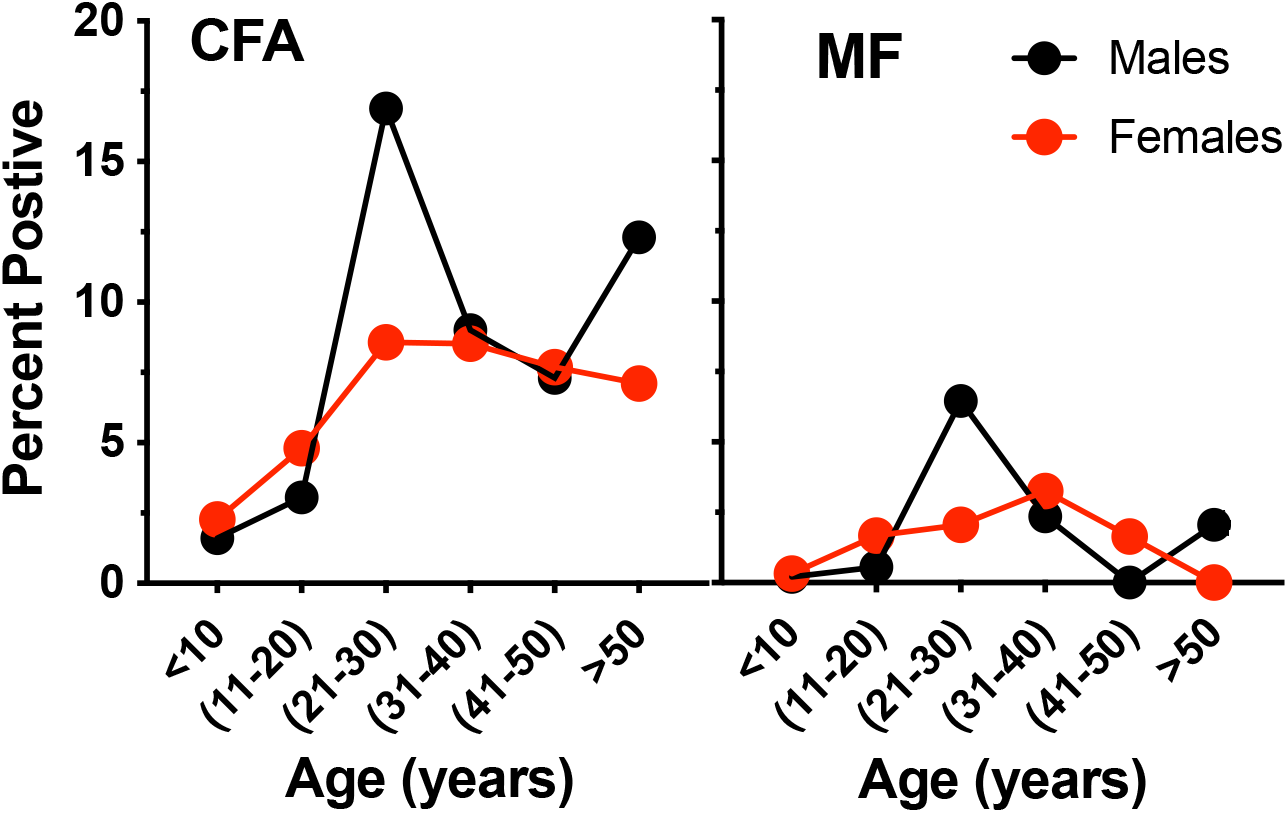
Circulating filarial antigen (CFA) and microfilaremia (MF) prevalence by age groups pre-MDA for 49 clusters.

**Table 2.** Study population demographic and infection characteristics of 6-9 and ≥ 10 years before MDA.

**a. 6-9 years**
| *District* | *\*Total population* | *Villages surveyed* | *N* | *Percent Females* | *CFA+ (N)* | *CFA % (95%CI) [range]* | *Mf+ (N)* | *Mf % (95%CI) [range]* | *Villages ≥2% CFA* | *Villages ≥1% Mf* |
| --- | --- | --- | --- | --- | --- | --- | --- | --- | --- | --- |
| Kokopo | 10,939 | 11 | **480** | 47 | 23 | 4.8 (3.1-7.1) [0-21.6] | 5 | 1.0 (0.3-2.4) [0-7.8] | 3 | 2 |
| Gazelle | 20,396 | 17 | **660** | 51 | 10 | 1.5 (0.7-2.8) [0-6.0] | 0 | - | 5 | - |
| Pomio | 10,437 | 17 | **581** | 45 | 4 | 0.7 (0.2-1.8) [0-7.2] | 0 | - | 3 | - |
| Rabaul | 3,415 | 4 | **185** | 52 | 0 | - | 0 | - | - | - |
| **Total** | **45,188** | **49** | **1,906** | **48** | **37** | **1.9 (1.4-2.7) [0-21.6]** | **5** | **0.3 (0.1-0.6) [0-7.8]** | **11** | **2** |

\*The estimated population of 6-9-year-olds in ENBP according to the population pyramid rate for PNG.

**b. $\geq 10$ years**
| *District* | *\*Total population* | *Villages surveyed* | *N* | *Percent Females* | *CFA+ (N)* | *CFA % (95%CI) [range]* | *Mf+ (N)* | *Mf % [range]* | *Villages $\geq 2\%$ CFA* | *Villages $\geq 1\%$ Mf* |
| --- | --- | --- | --- | --- | --- | --- | --- | --- | --- | --- |
| Kokopo | 69,284 | 11 | 477 | 66 | 46 | 9.6 (7.2-12.7) [0-52.0] | 24 | 5.0 (3.3-7.4) [0-36.0] | 6 | 3 |
| Gazelle | 129,174 | 17 | 827 | 58 | 30 | 3.6 (2.5-5.1) [0-15.7] | 6 | 0.7 (0.3-1.6) [0-6.1] | 8 | 3 |
| Pomio | 66,104 | 17 | 827 | 62 | 96 | 11.6 (9.5-14.0) [0-36.8] | 16 | 1.9 (1.1-3.1) [0-5.8] | 12 | 9 |
| Rabaul | 21,627 | 4 | 215 | 64 | 4 | 1.9 (0.5-4.7) [0-4.9] | 1 | 0.5 (0.0-2.6) [0-2.4] | 2 | 1 |
| **Total** | **286,190** | **49** | **2,346** | **61** | **176** | **7.5 (6.5-8.6) [0-52.0]** | **47** | **2.0 (1.5-2.7) [0-36.0]** | **28** | **16** |

\*The estimated population of $\geq 10$ -year-olds in ENB according to the population pyramid rate for PNG.

LF infection in ENBP was highly heterogeneous, with the highest prevalence concentrated in the Duke of York Islands (DOY) in the Kokopo district and Wide Bay Area along the eastern coast of the Pomio District (**Figure 3**). Gazelle and Rabaul Districts had lower LF infection prevalence.

**Figure 3.**
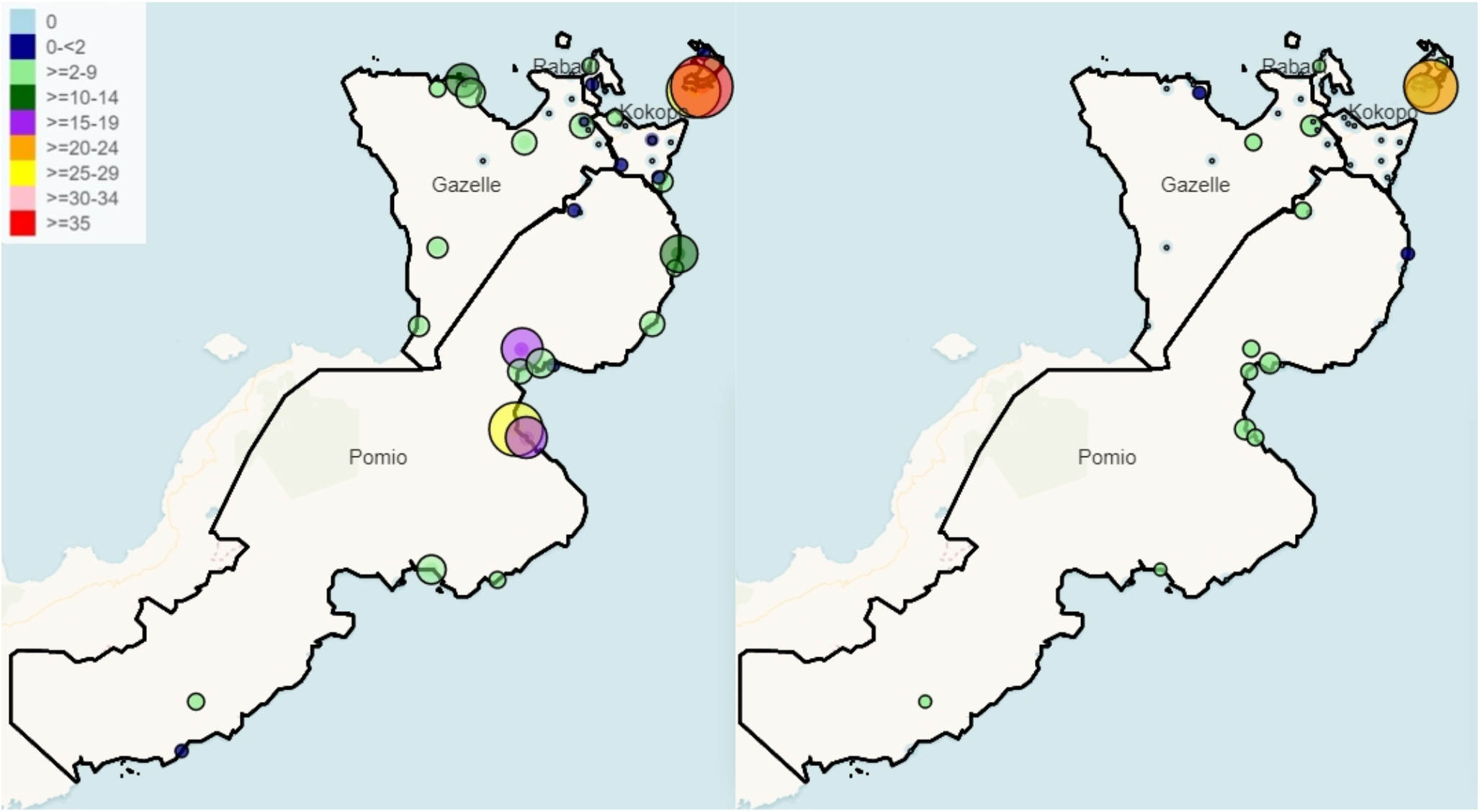
Spatial distribution of lymphatic filariasis circulating antigen positivity (CFA, left panel) and microfilaremia rates (Mf, right panel) throughout ENBP before mass drug distribution. Small solid dots indicated communities without any detectable LF infection. The colors and circle size represent LF prevalence for CFA or Mf.

### Effect of sampling design for detecting LF infected villages

Sampling protocols affected the efficiency of detecting LF infection. In the 30 villages sampled by PPS, 2.3% (95% CI 1.8-2.9%; 59/2,561) were CFA positive, and 0.6% (95% CI 0.3-0.9%; 14/2,561) were Mf positive. In children 6-9 years of age in the 30 villages selected by PPS, only 0.7% (95% CI 0.3-1.4; 8/1,121) were CFA positive, and none were Mf positive. Nine of 30 (30%) PPS selected villages had CFA >2%. (**Figure 5 and Supplemental Table 4**). Infection in children and adults in the purposively selected villages was higher than in randomly selected villages (**Figure 5, Supplemental Table 5**). The five original purposively selected villages had >2% CFA positive, and 3 had Mf prevalences of >1% (**Figure 5**). Of the additional 14 villages purposely selected in rural villages, 10 (71.4%) villages had χ′ 2% CFA, and 4 of 14 (28.6%) had >1% Mf prevalence.

**Figure 5.**
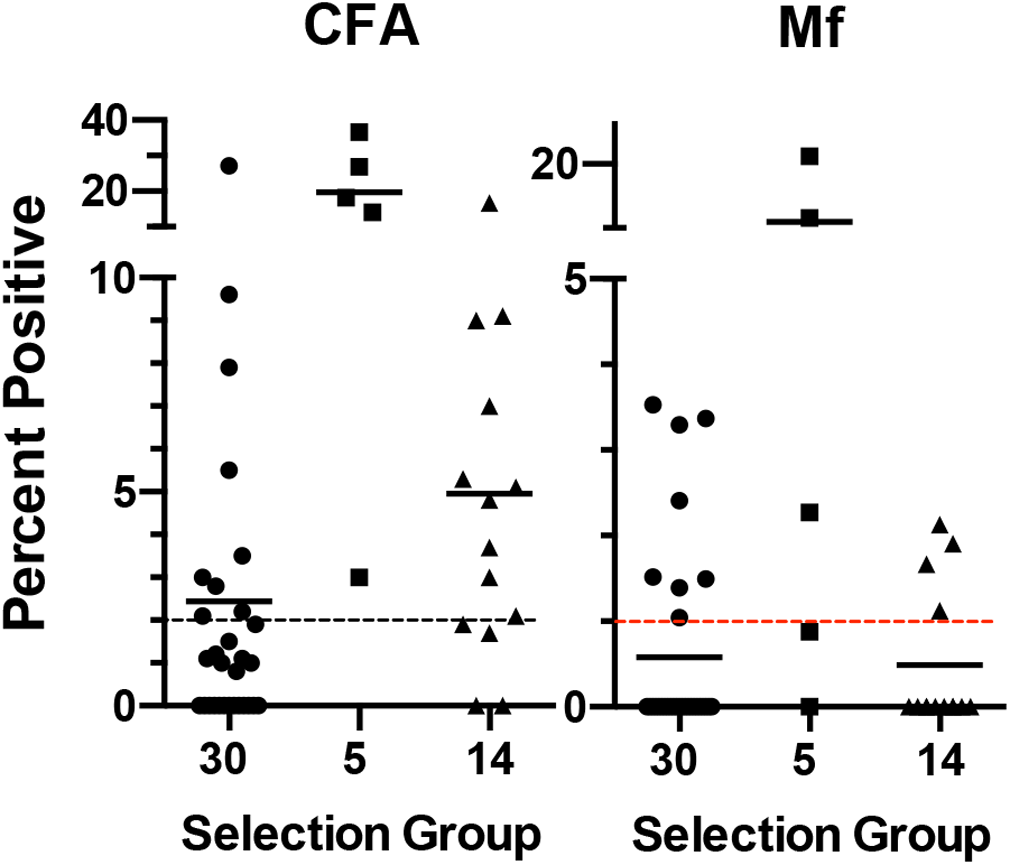
LF infection parameters are shown according to different sampling strategies for cluster selection. Thirty clusters used a population proportion random sample. ENBP Provincial Health Authority purposefully selected five additional clusters with prior evidence of LF. Fourteen more clusters were chosen to adequately sample rural areas in Pomio (N=9) and Gazelle (N=5) districts. The dashed line on the left panel and the red line on the right panel represent WHO-specified thresholds of 2% CFA and 1% MF rates, suggesting ongoing transmission in the cluster if above these thresholds.

### Mass drug administration

The first round MDA was administered by the ENBP Provincial Health Authority in November 2019 with the support PNG National Department of Health and Japan International Cooperative Agency (JICA) to 307,568 of 376,566 total residents of ENBP (81.7%, reported or administrative coverage), **Supplemental Table 6**). The reported coverage varied by district; from rural districts of Pomio (72.8%) and Gazelle (83.8%) to 93.3% and 96.7% coverage in the more urban districts of Kokopo and Rabaul, respectively. Of note, only DEC plus Albendazole was administered to children 2 to 4. In December 2019, an independent coverage survey was performed to validate the administrative coverage survey. This included 2,598 individuals in 450 randomly selected households in 45 randomly selected villages. Over 80% of the individuals reported taking treatment in all age groups except those >50 years of age (**supplemental Figure 1**). Reported MDA compliance was equivalent in males and females and >80% in males 21-30 years (with the highest baseline LF prevalence). The most common reason for not taking the drugs was absenteeism in all 4 districts. Residents of Rabaul also reported insufficient LF awareness and incorrect age classification, which sometimes led to the exclusion of individuals > 55 years of age.

### Model-Based geospatial sampling post-MDA LF infection parameters

Because the pre-MDA population proportionate sampling oversampled urban areas with lower LF prevalence and under-sampled rural areas with higher LF prevalence, we undertook a sampling strategy to reflect better the geographical distribution of LF in ENBP. Thus, we used model-based geospatial (MBG) as an alternative method to select clusters for monitoring and evaluation during the post-MDA 1-year follow-up survey. Circulating filarial antigen (CFA) prevalence data from the baseline survey was used to assess the risk probability for LF in ENBP (**Figure 6**). Besides location, the model considered the following covariates: relative humidity (RH), annual precipitation (AP), annual temperature (AT), elevation (E), distance from the sea (DS), and socio-economic status (SEI). Because of the small sample size, we combined the covariates into a single covariate using principal component analysis (PCA) and used the first component (PCA1) as a predictor for LF. More specifically, RH - 0.053, AP - 0.502 gave this AT + 0.502 E+ 0.502 DS and explained about 62% of the total variation in the environmental variables. The sample size was too small to establish which variables were most informative for predicting areas with >2% CFA. A hotspot area with an estimated prevalence above >2% is identified in the central part of the eastern coast and in Duke of York Islands. The model selected 50 villages for sampling across the four districts of ENBP that were likely to have >2% of CFA positivity using baseline LF data and co-variates associated with the presence of LF. Among the 50 sites, the model captured all 24 baseline clusters with >2% CFA and selected 26 not sampled in the baseline survey.

**Figure 6.**
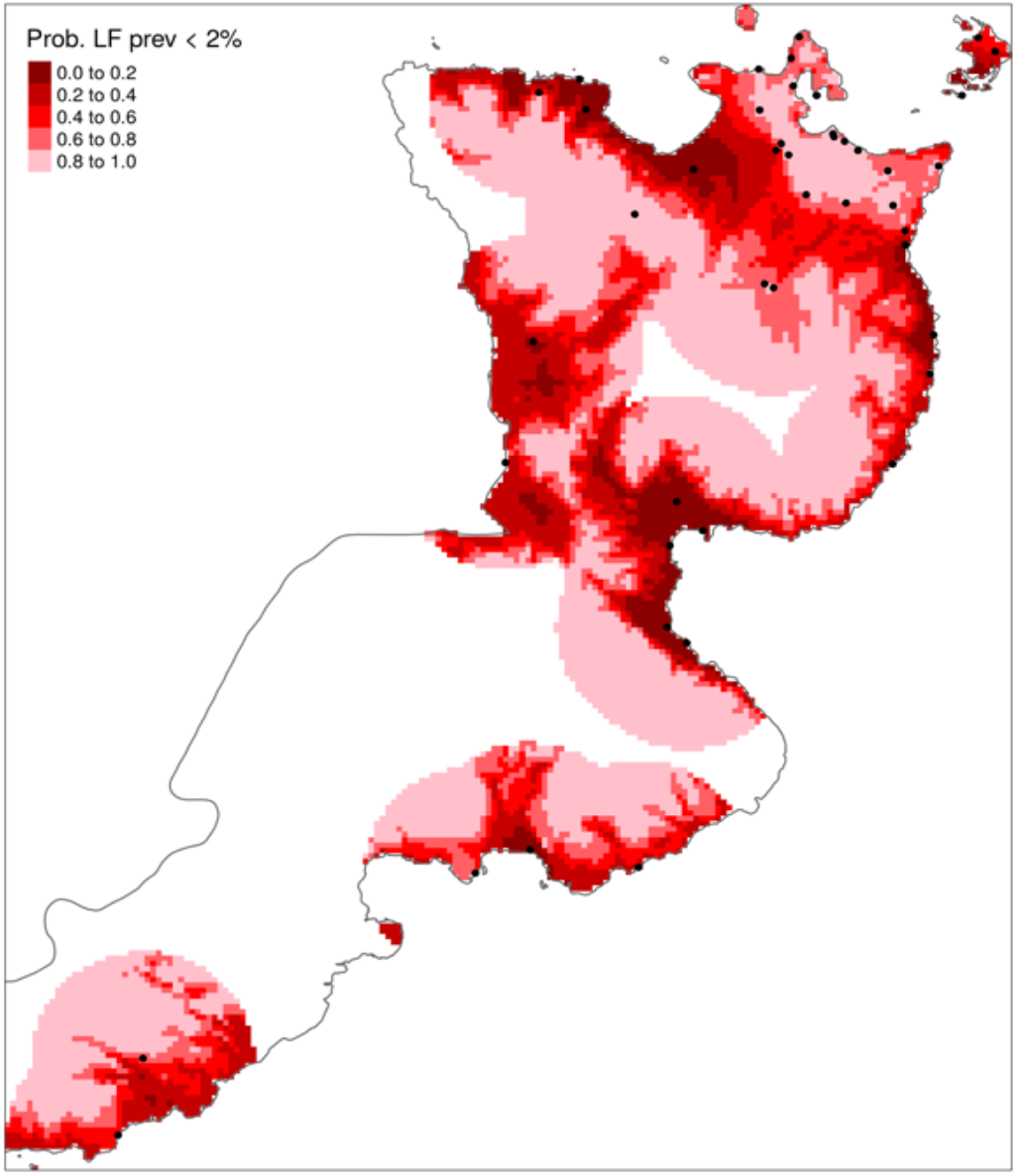
Geospatial probability map of LF prevalence. The dark red area has a low probability that CFA prevalence is <2%; thus, these areas are more likely to have LF. The map resolution is 1 km. White areas have insufficient data to assess LF risk. Risk mapping was generated based on covariates of elevation, rainfall, temperature, distance from the sea, socioeconomic status, and LF prevalence using baseline survey clusters (black dots).

Of the 50 clusters selected, 47 were surveyed, with 4,611 participants from 6-9 and ≥ 18-year-old age groups, to capture Mf-positive individuals (**Table 1**) better. Geostatistical modeling selected the 24 villages with >2% CFA identified pre-MDA (**Table 1, supplemental Table 7)**. Following MDA-IDA, 20 of 23 newly selected villages had >2% CFA, indicating an efficient selection of communities using MBG. Children were surveyed in 19 of the 47 clusters, and adults (≥18 years) were studied in all 47. Because of the differences in sampling design pre- and post-MDA, we focused on changes in LF infection parameters in age-matched groups in the 24 clusters monitored pre- and post-MDA in children and adults. In the 24 sentinel sites matched pre- and post-MDA for individuals ≥18 years of age, the CFA mean prevalence decreased by 38.6% from 17.3% (95% CI 16.2-18.4; 141/816) to 10.6% (95% CI 10.1-11.2; 215/2,015; P<0.0001). By contrast, Mf prevalence decreased by 90.1% from 4.9% (95% CI 4.28-5.52; 40/816) to 0.45% (95%CI 0.2-0.8; 9/2016, P<0.0001) (**Figure 7**). Eleven of the 24 villages had children 6-9 years with LF infection before MDA, and 6-9 years old were re-evaluated in these same villages post-MDA. CFA prevalence decreased by 30.1% from a CFA prevalence of 7.2% (95% CI 4.8-10.5; 30/417) to 4.9% (95% CI 3.0-7.7; 18/365, P=0.23). MDA completely cleared Mf infection in children from a prevalence of 1.2% (95% CI 0.4-2.7; 5/417) to 0% (95% CI 0.0-1.0; 0/365; P=0.065).

**Figure 7.**
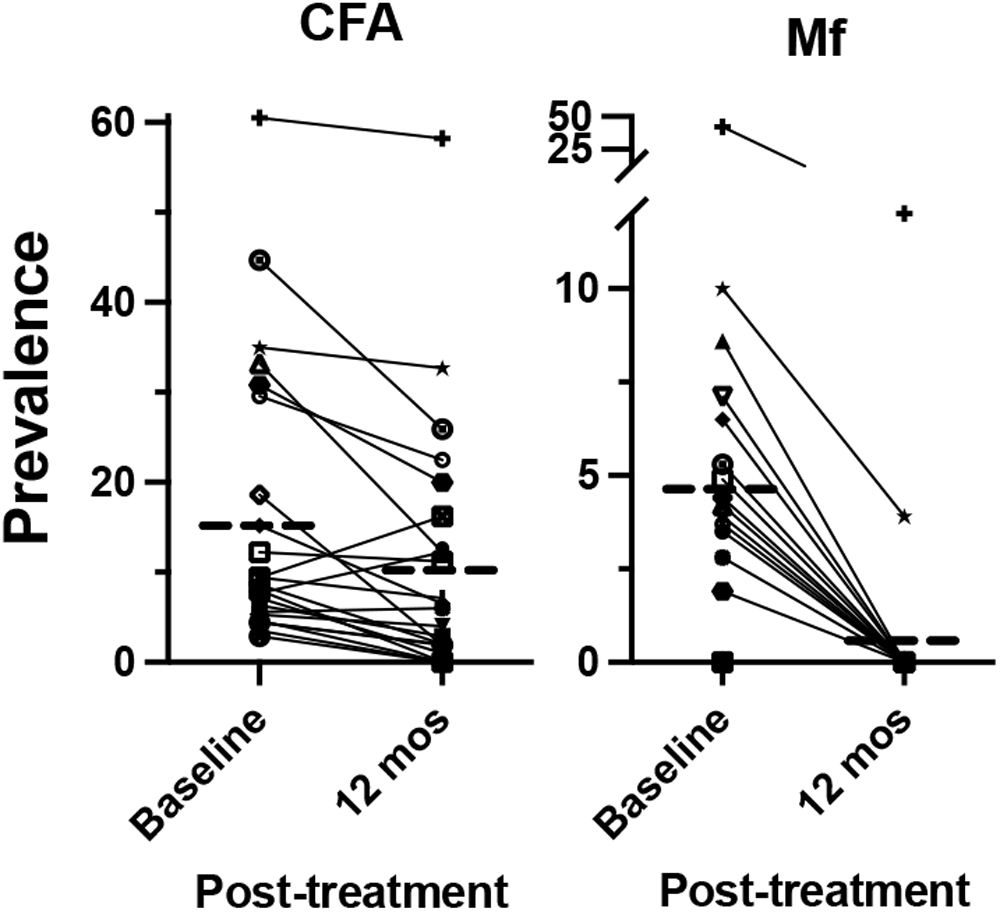
Impact of treatment on CFA and Mf prevalence in 24 clusters before and after MDA. Individuals shown are ≥18 years old with N=816 before MDA and N=2015 12 months post-MDA. Dashed lines represent means.

Children 6-9 years in 8 additional villages were examined for LF infection parameters post-MDA combined with 11 villages described above for a total 19. Post-MDA CFA prevalence in children 6-9 years (in 19 villages) was 2.9% (95% CI 1.8-4.2; 20/694) was comparable to 1.9% (95% CI 1.4-2.7; 37/1,906; P=0.17) among children pre-MDA (**Table 2a**), further demonstrating little change in CFA prevalence one-year post-MDA. By contrast, no Mf-positive children were identified post-MDA (0/694) compared to 0.3% pre-MDA (5/1,906; **Table 2a**).

Overall, post-MDA-IDA ≥ 18-year-old CFA prevalence of 9.0% (95% CI 8.1-10.0; 352/3,916, **Table 3b**) was comparable to pre-MDA CFA prevalence among the same age group of ≥18 years of 9.6% (95% CI 8.2-11.1; 151/1,581; **supplemental table 2e**). By contrast, MDA-IDA markedly reduced overall Mf prevalence to 0.33% (95% CI 0.21-0.42; 15/4611). Two districts, Rabaul and Gazelle, had no sentinel villages with ≥1% Mf positivity. However, 6 sentinel villages had ≥1% Mf. Three clusters ≥1% Mf occurred in the Duke of York Islands (Kabatira, Karawara, and Utuan, 1%, 2% and 7% respectively) and the remainder of the sentinel sites in the Kokopo district were Mf negative (**Figure 8**). The other 3 clusters with 21% Mf positivity post-MDA were clustered along the East Coast in the Wide Bay area of the Pomio district, areas that also had high baseline LF prevalence.

**Figure 8.**
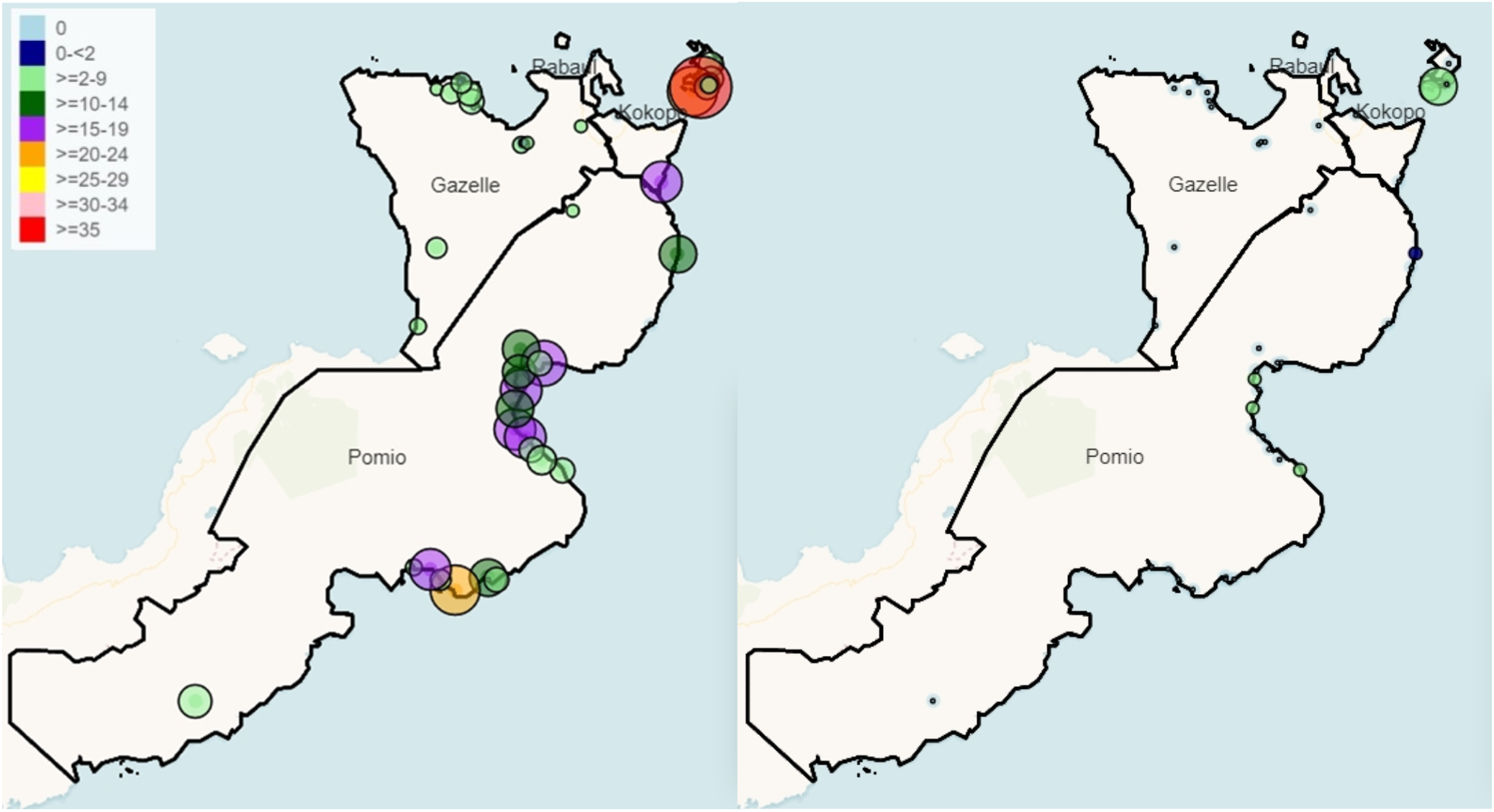
Geographic distribution and prevalence of clusters sampled following MDA. The left panel shows CFA prevalence, and the right panel shows Mf prevalence. Small white dots are clusters without detectable CFA or Mf in participants. The color gradient of the maps corresponds to the different CFA prevalence (left panel). The size of green circles correlates with Mf prevalence, with the smallest green circles being 1% Mf.

**Table 3.** Study population demographic and infection characteristics of 6-9 and ≥ 18 years at 1 year-post MDA.

**a. 6-9 years**
| *District* | *Villages surveyed* | *N* | *Percent Females* | *CFA+ (N)* | *CFA % (95%CI) [range]* | *Mf+ (N) [range]* | *Mf %* | *Villages ≥2% CFA* | *Villages ≥1% Mf* |
| --- | --- | --- | --- | --- | --- | --- | --- | --- | --- |
| Kokopo | 5 | 231 | 50.7 | 9 | 3.9 (1.8-7.3) [0-10.6] | 0 | - | 2 | 0 |
| Gazelle | 2 | 93 | 52.9 | 1 | 1.1 (0.0-5.9) | 0 | - | 1 | 0 |
| Pomio | 12 | 370 | 46.6 | 10 | 2.7 (1.3-4.9) | 0 | - | 5 | 0 |
| Rabaul | 0 | 0 | - | - | - | - | - | - | - |
| **Total** | **19** | **694** | **48.8** | **20** | **2.9 (1.8-4.4)** | **0** |  | **8** | **0** |

**b. ≥18 years**
| *District* | *Villages surveyed* | *N* | *Percent Females* | *CFA+ (N)* | *CFA % (95%CI) [range]* | *Mf+ (N)* | *Mf % [range]* | *Villages ≥2% CFA* | *Villages ≥1% Mf* |
| --- | --- | --- | --- | --- | --- | --- | --- | --- | --- |
| Kokopo | 9 | 627 | 64.9 | 91 | 14.5 (11.9-17.6) [0-58.2] | 10 | 1.6 (0.8-2.9) [0-12.7] | 8 | 3 |
| Gazelle | 12 | 1,107 | 54.2 | 34 | 3.1 (2.1-4.3) [0-18.7] | 0 | 0.1 (0.0-0.1) [0.9] | 9 | 0 |
| Pomio | 25 | 2,083 | 55.8 | 227 | 10.9 (9.6-12.4) [0-28.4] | 5 | 0.3 (0.1-0.5) [0.9-1.0] | 22 | 3 |
| Rabaul | 1 | 99 | 57.6 | 0 | 0 | 0 | 0 | 0 | 0 |
| **Total** | **47** | **3,916** | **56.84** | **352** | **9.0 (8.1-10.0) [0-58.2]** | **15** | **0.4 (0.2-0.6) [0-12.7]** | **39** | **6** |

### Age-specific impact of MDA on LF infection parameters: 6-9 and ≥18-year age-groups

Out of the 4,611 participants surveyed at 1-year post-MDA, 694 were aged 6-9 years and 3916 were adults (≥18 years). Children (6-9 years) were surveyed in 19 of the 47 clusters and adults (218 years) were surveyed in all 47. The CFA prevalence in the children group was 2.88% (95% CI 1.77-4.42; 20/694) with no Mf infection (**Table 2a**). Eight clusters of which children were surveyed exceeded the 2% CFA threshold. In the adult population, the CFA prevalence was 9.01% (95% CI 8.13-9.95; 352/3916) and 0.36% (95% CI 0.20-0.60; 14/3916) mf positivity **(Table 2b**). Thirty-nine of the 47 clusters of which adults were surveyed had CFA prevalence of ≥2% and 3 clusters with Mf prevalence of ≥1%.

## Discussion

East New Britain was the first treatment province in PNG to implement MDA with IDA. The program was highly successful and achieved >80% drug coverage after the first round of MDA. Following one round of MDA, overall Mf prevalence decreased from 1.2% pre-MDA to 0.3%, well below the WHO pre-TAS Mf prevalence target of <1%. However, some areas with high prevalence at baseline failed to reach this target. One of the four districts in the province detected no Mf-positive individuals, and another district one Mf-positive individual following MDA-IDA, suggesting little risk of ongoing LF transmission. By contrast, 6 of 47 villages surveyed had post-MDA Mf prevalence 21%, indicating ongoing risk for LF transmission in these areas. These remaining Mf-positive communities were clustered in small geographic areas with high LF prevalence pre-MDA. Current WHO guidelines recommend another round of MDA for the whole province [14], which has the advantage of treating other parasitic infections or capturing Mf carriers from nearby unsampled villages.

This study highlights the strengths and weaknesses of different sampling strategies and endpoints to assess LF prevalence where infections are highly focal and prior information on LF is limited. The population proportional sampling strategy (PPS, WHO recommended) used at baseline underestimated the LF burden. This is because PPS oversampled communities in urban areas with little LF and under sampled communities in rural areas with more LF. Results from this study also add to recent reports that highlight the limitations of TAS of young school-aged children for post-MDA stopping decisions. Before MDA, only 0.7% of children 6-9 years of age in ENBP were CFA positive, and 0.6% among children 6-7 years old, the TAS target population, which is well below the 2% threshold believed to suggest recent or ongoing LF transmission. Indeed, ENBP probably could have passed TAS performed according to WHO protocols before any MDA, even though the province had areas with high endemicity at that time. Since only a few infections in young children were detected before MDA, TAS would have provided unreliable information for MDA-stopping decisions following MDA. Despite low infection prevalence in children, ENBP was known to be endemic for LF based on prior surveys conducted by health officials and confirmed by baseline surveys reported here. However, many other areas in PNG do not have detailed information on LF prevalence because LF testing and reporting are not included in the country’s National Health Information Surveillance System. Our results from ENBP suggest that PPS and TAS may not be the best way forward for LF elimination efforts in other regions in PNG.

This study reinforced the difficulty of using CFA alone as an endpoint for assessing the impact of IDA on LF transmission. One round of MDA reduced CFA prevalence by 30.1% in children 6-9 years of age and 38.6% in adults; this lower CFA reduction in adults has been observed in prior clinical trials [8, 9, 11–13]. However, worms that survive IDA treatment can continue to release CFA for years after Mf clearance [9]. One approach to this problem for MDA-stopping decisions and post-MDA surveillance is to use CFA as a screening test in adults, with Mf testing restricted to those with positive antigen tests. It should be safe to stop MDA if Mf prevalence has been reduced to very low levels in adults despite CFA prevalence >2%. Mosquitoes cannot transmit LF without access to Mf.

We used geospatial modeling to more efficiently identify communities at risk for LF and assess the impact of one round of MDA. Geospatial modeling is increasingly used for mapping tropical diseases in low-resource settings where disease distribution data are limited or non-existent [21]. It incorporates information on known environmental and sociodemographic risk factors for LF, including elevation, vegetation, and population density. It has been increasingly used to map tropical diseases, such as malaria, Chagas disease, LF, and trachoma [22–24] [25]. The method has also been applied to predict residual LF hotspots post-MDA. Our results show that geospatial modeling was more helpful than PPS for identifying communities with high CFA post-MDA using baseline infection data and environmental variables. By facilitating the detection of persistent infections, this approach should help inform programmatic decisions [26]. Because of the small sample size, the analysis could not identify individual factors in the model most predictive of LF infection.

Prior WHO recommendations that stopping decisions for MDA based on the mean prevalence in an EU is insufficient. As LF transmission approaches interruption, the remaining LF infection becomes increasingly focal, and these foci should be targeted for possible further MDA-IDA. This study highlights the concept. One round of MDA-IDA with high coverage likely eliminated LF transmission in two of four districts in ENBP with low baseline LF prevalence. Additional rounds of MDA-IDA could target the two districts with clusters that have >1% Mf, or focus on clusters within the district, e.g. the Duke of York Islands. The WHO recommends two rounds of MDA-IDA, which may capture those who missed the first round of MDA and further treat other infections. However, in LF-endemic areas like PNG, where LF infection may be highly focal, MDA costly, a single round of MDA-IDA that achieves >80% coverage, the IDA impact survey could occur after the first round of MDA. This would allow additional rounds of MDA-IDA to target areas with >1% Mf prevalence.

Several limitations should be mentioned regarding this study. Before MDA, we sampled many children so that the median age of our sample was 12 years. In contrast, our post-MDA survey focused more on adults (median age of 29 years). Since LF is age-dependent, a direct comparison of infection prevalence before and after MDA required a comparison of an age-matched subset of individuals sampled before and after MDA. Another limitation of the study is that it only assessed the impact of one round of MDA, although WHO recommends two rounds. This was intentional because we wanted to determine whether a single round of MDA with high coverage would reduce Mf prevalence to <1%. This did occur in many areas, but hotspots remained.

In conclusion, this study showed that a single round of MDA-IDA effectively reduced Mf prevalence to very low levels in most endemic localities in ENBP. This was due to the excellent efficacy of IDA in clearing Mf in PNG and the high MDA coverage provided by public health officials in ENBP. Additional surveys will be needed to determine whether a second round of MDA-IDA has been sufficient to clear up remaining hot spots of infection. Results from this study will support the expanded use of IDA for LF elimination in logistically challenging areas with significant resource limitations. They also clearly demonstrated the importance of sampling adults rather than young children post-MDA and the value of geospatial modeling for identifying high risk locations better to target monitoring and evaluation efforts for LF elimination programs.

## Data Availability

Detailed village specific data is included in the supplemental information

## Acknowledgments

We thank the staff of the East New Britain Provincial Health Authority and Papua New Guinea National Department of Health. In addition, we thank the teams who undertook the sample collections. We also thank the regional health directors, village leaders, and especially our study participants. The Bill & Melinda Gates Foundation (OPPGH5342 to GW and PF) funded the project.

## Contributors

KB, BM, CB, DT, MS, GW, PF, LR, ML, and CK were responsible for the study design. KB performed a literature search. KB, CB, MP, DT, and MS collected data; KB, DT, JK, EG, GW, PD, and CK analyzed and interpreted the data. KB and CK wrote the manuscript, which was revised by GW and PF.

